# Novel Coronavirus 2019 (Covid-19) epidemic scale estimation: topological network-based infection dynamics model

**DOI:** 10.1101/2020.02.20.20023572

**Authors:** Keke Tang, Yining Huang, Meilian Chen

## Abstract

**Backgrounds:** An ongoing outbreak of novel coronavirus pneumonia (Covid-19) hit Wuhan and hundreds of cities, 29 territories in global. We present a method for scale estimation in dynamic while most of the researchers used static parameters.

**Methods:** We use historical data and SEIR model for important parameters assumption. And according to the time line, we use dynamic parameters for infection topology network building. Also, the migration data is used for Non-Wuhan area estimation which can be cross validated for Wuhan model. All data are from public.

**Results:** The estimated number of infections is 61,596 (95%CI: 58,344.02-64,847.98) by 25 Jan in Wuhan. And the estimation number of the imported cases from Wuhan of Guangzhou was 170 (95%CI: 161.27-179.26), infections scale in Guangzhou is 315 (95%CI: 109.20-520.79), while the imported cases is 168 and the infections scale is 339 published by authority.

**Conclusions:** Using dynamic network model and dynamic parameters for different time periods is an effective way for infections scale modeling.

## Introduction

Multiple similar pneumonia cases of unknown aetiology were identified by authority in Wuhan, China (Tan et al.,2020). According to the report, the first case was appeared on 1 December, 2019. By Jan 2, 2020, there were total 41 patients who had been identified as having laboratory confirmed 2019-nCoV infection, and 66% of them had Huanan Seafood Wholesale Market exposure (Huang et al.,2020). And by Jan 11, the confirmed cases had a sharp rise to 248 (Li et al.,2020). On 21 January 2020, the WHO suggested there was possible sustained human-to-human transmission after renowned scientist Nanshan Zhong made this message to public. And on 23 January 2020 a quarantine on travelling in and out of Wuhan was executed. It’s noticed that Jan 10 was the start of Chinese New Year migration period and Wuhan was a major transport hub of the country which owned one of the four most important railway stations in China, which catalyzed the wide spread of whole country and global. From Jan 23, there were many responses executed in domestic, including similar quarantine measures in multiple cities of Hubei and Non-Hubei cities, medical aid, identified and suspected cases tracking. Fortunately, they made progress proved by the constantly going down of confirmed cases.

As of 1000 GMT 18 Feb 2020, 73,424 cases had been confirmed in 26 countries, and 98.66% of them came from China mainland. It’s noticed that some territories like Japan the confirmed cases is rising up, although the MHLW of Japan have no confirmed breakout in domestic. It’s possible that these territories may in early stage of breakout and it’s believed that the data from China, especially Wuhan region may contribute to infection control. Some researchers have published report on infection scale estimation at difference time points. Wu estimated that the basic reproductive number for 2019-nCoV was 2.68 (95% CI 2.47–2.86) and 75,815 individuals (95% CI 37,304-130,330) had been infected in Wuhan as of Jan 25, 2020 (Wu et al.,2020). Read considered that 21,022 (11,090–33,490) total infections in Wuhan from 1 to 22 January (Read et al.,2020). However, the estimation of these studies basically focused on merely one time point and haven’t considered the parameter changing after quarantined.

In this article, we propose an estimation model base on topological network and try to make estimation of infection scale of Wuhan. The quarantined factor is included into consideration within different time lines. Further more, We also use data of a Non-Hubei city, Guangzhou, for cross validation estimated by migration scale.

## Method

### Assumptions of parameters

The equations of R_0_ based on SEIR model are as follows:

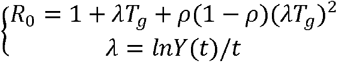

Where t is the outbreak time of the disease, T_g_ is the generation time, ρ is the confirmed rate of suspected cases, and Y_(t)_ is the actual infection number of t days of the disease. According to Li’s study on early cases, many cases of “unknown pneumonia” began to appear in mid-December. Specifically, there were 11 cases from December 10 to December 20 (Li et al.,2020). Meanwhile, the earliest confirmed case appeared on December 8 and there was no history of contact with the Huanan Seafood Market. Therefore, based on the average incubation stage of 5.2 days (the incubation stage for 95% of confirmed patients is about 12.5 days), the situation of human-to-human transmission took place from mid-December. The specific time can not be estimated. As a result, we assume that the outbreak started on December 10. At present, no satisfactory parameter evidence has been found with respect to the confirmed rate ρ of suspected cases. Based on the historical data of new suspected cases on the t day and new confirmed cases on the t + 1 day (it takes 10-12 hours for the current detection technology to get results), and based on the Yang’s retrospect study of 8,866 cases, the confirmed rate of suspected cases is about 46% (Yang et al.,2020). With reference to the historical data of SARS (Chowell et al.,2020), T_g_ is 8.4 days. Based on the assumption above, R_0_ can be calculated. (Table 1)

**Table 1.**
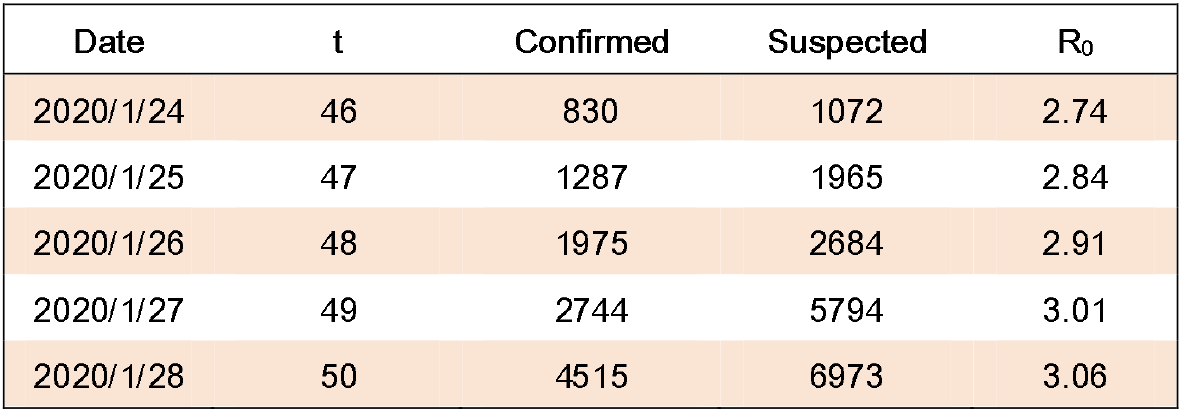
R_0_ calculation by the data from Jan 24 to Jan 28 made public by National Health Commission of the People’s Republic of China.

The basic reproduction rate can be defined as:

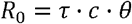

Where *τ* is the probability of infection, *c* is the contact rate between the susceptible and the infected and *θ* is the infection cycle.

For simplicity, estimates can be made based on the SIR model. Suppose N is the population base, S is the susceptible number of populations, I is the number of infections, and R is the number of removals (including cure and death, assuming that cure will not be infected again).

Therefore, s=S/N,i=I/N and r=R/N stand for the ratio of different crowds. Based on model SIR, we have:

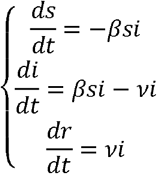

*β* = *τ* · *c* and *v* are constants, then *θ = v*^-1^. When at the starting point of the outbreak:

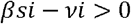

So:

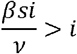

It’s considered that the whole population is susceptible, then s=1:

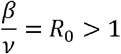

Based on the data so far, we have v=0.06, then β=0.18.

### Topology network

Many complex systems in the real world can be represented in the form of networks. For example, if a person is regarded as a node in the network, then the connection between people can be considered as an edge connecting nodes. That is a way which a social relationship network can be constructed in. Similarly, in epidemiology, individuals are nodes, and the contacts of individuals can be seen as edges. Moreover, the state of individual (such as susceptibility, exposure, and infection) can be added to nodes as additional information. As a result, an epidemiological network topology graph can be constructed with these elements.

One of the most important parameters is the probability of contact between people. For example, a person living in the city (work, transportation, dining) may need to contact dozens of people in close range, and indirectly contact hundreds or even thousands (such as in the subway environment) in one day. If we assume that an individual contacts 1,000 people in Guangzhou every day, therefore, the probability of contact is 10/1.5×10^7^. This contact probability can be regarded as the generation probability of edges between nodes. We generate random network topology graphs by using the parameters mentioned above for presenting relationships among the crowd intuitively (Figure 1). The results of population isolation caused by quarantine measures for epidemic control are very similar with it. Therefore, we can generate a network topology graph based on the timeline of the epidemic in subsequent simulations by considering the contact situation of the crowd, which can reflect different situations more realistically.

**Figure 1.**
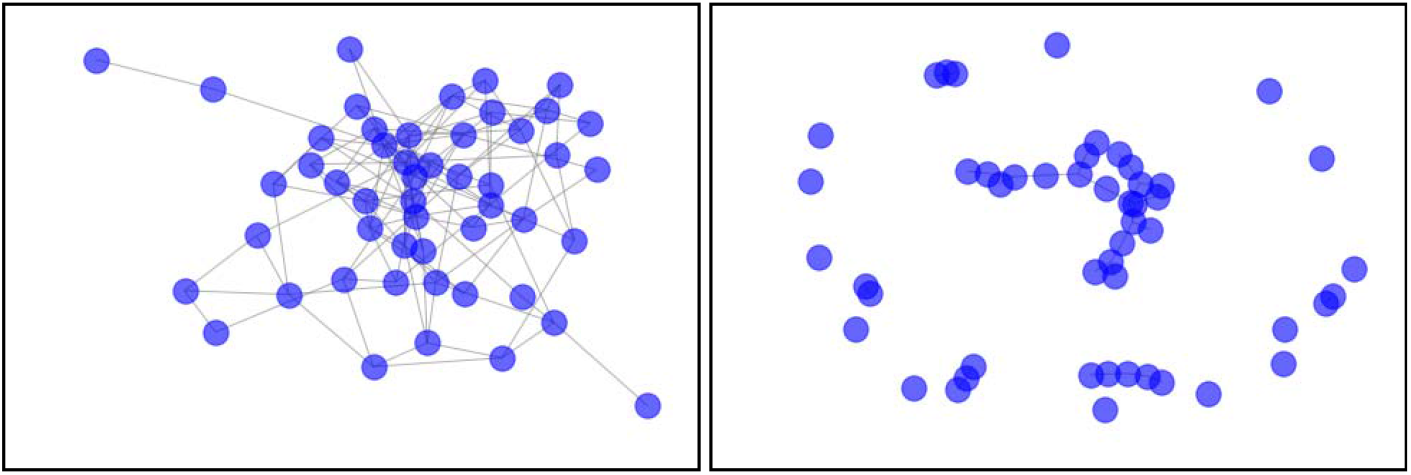
The left shows 50 nodes with a connection probability of 10%; And the right figure shows 50 nodes with a connection probability of 3%. It is obvious that there are some isolated communities on the right.

### Time line

According to the existing data, the overall time line of the NCP epidemic situation is sorted out. The purpose of that is to re-evaluate the important parameters of the model in different periods of time, so as to simulate the real situation as much as possible and make the prediction model more accurate. There are two important parameters that will be adjusted in different periods. The first one is contact rate(C). The second one is the propagation risk which could be directly adjusted by using the beta value mentioned above.

The time period can be divided into four parts: early breakout, development, spread and disease control stage. Due to the situation in Wuhan is relatively special, medical resources and suppliers were relative shortage within one week after quarantine. Fortunately, this situation was relieved by lager scale aiding after Feb 1. Considering that there may be a certain concentration in the hospital area, the contact rate may be higher than other areas.

## Result

### Scale estimation in Wuhan

#### 1. Early breakout, development and spread stage in Wuhan

An important parameter which should be taken into consideration is that how many actually infected people were at the start of the outbreak on December 10. According to Li ‘study, the number of confirmed cases as of December 10 was 7, while the number of confirmed cases as of December 31 was 47. If the maximum incubation period was 14 days, the actual number of infected cases on December 15 was close to 50. Besides, we assume that the contact rate C is 0.0001 as baseline, and infectivity was not excluded within the incubation period, and other parameters are the same with the analysis above. The estimation (Figure 3) shows that the number of infections experienced an increase since the end of December, and the number of new exposed had been accumulating until January 10, which led to the subsequent large scale of outbreak. In addition to considering the Spring Festival travel, this simulation is more in line with the argument that the “golden window period” of controlling the epidemic situation is before January 10. And after 18^th^ the infection scale underwent a sharp increase. The number of infections estimated on January 25 is showed as follows. The overall infection rate is about 0.684% and the 95% CI is 0.648%-0.721%. Because of the overall population of Wuhan during the Spring Festival is in a dynamic state, 9 million population as baseline is assumed. The number of infections is about 61,596, and the 95% CI is from 58,344.02 to 64,847.98.

**Figure 2.**
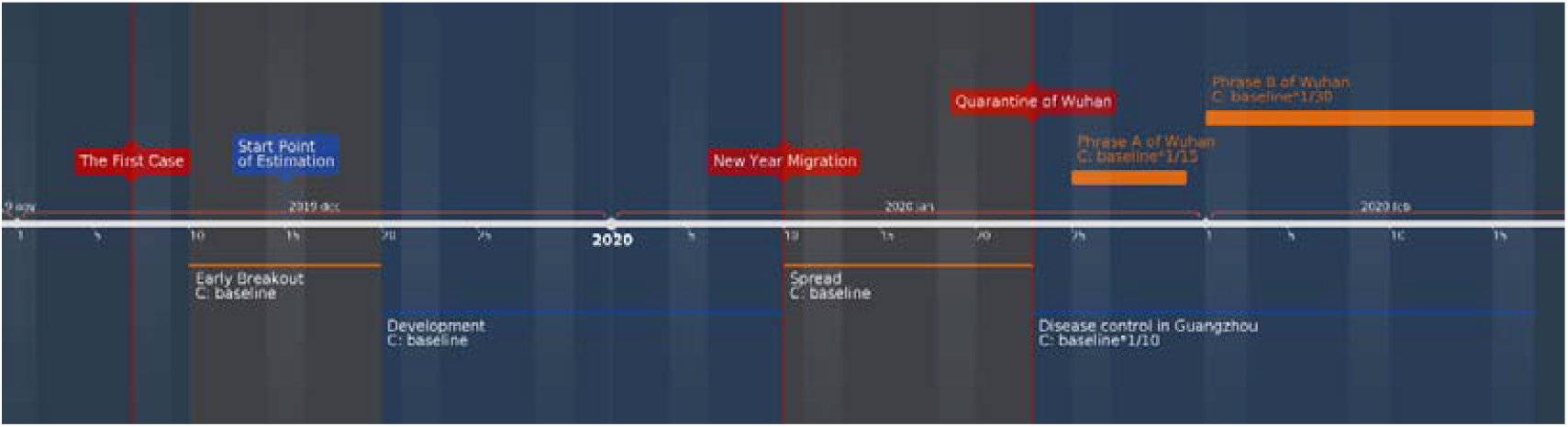
The timeline and Contact rate(C) assumed in different periods.

**Figure 3.**
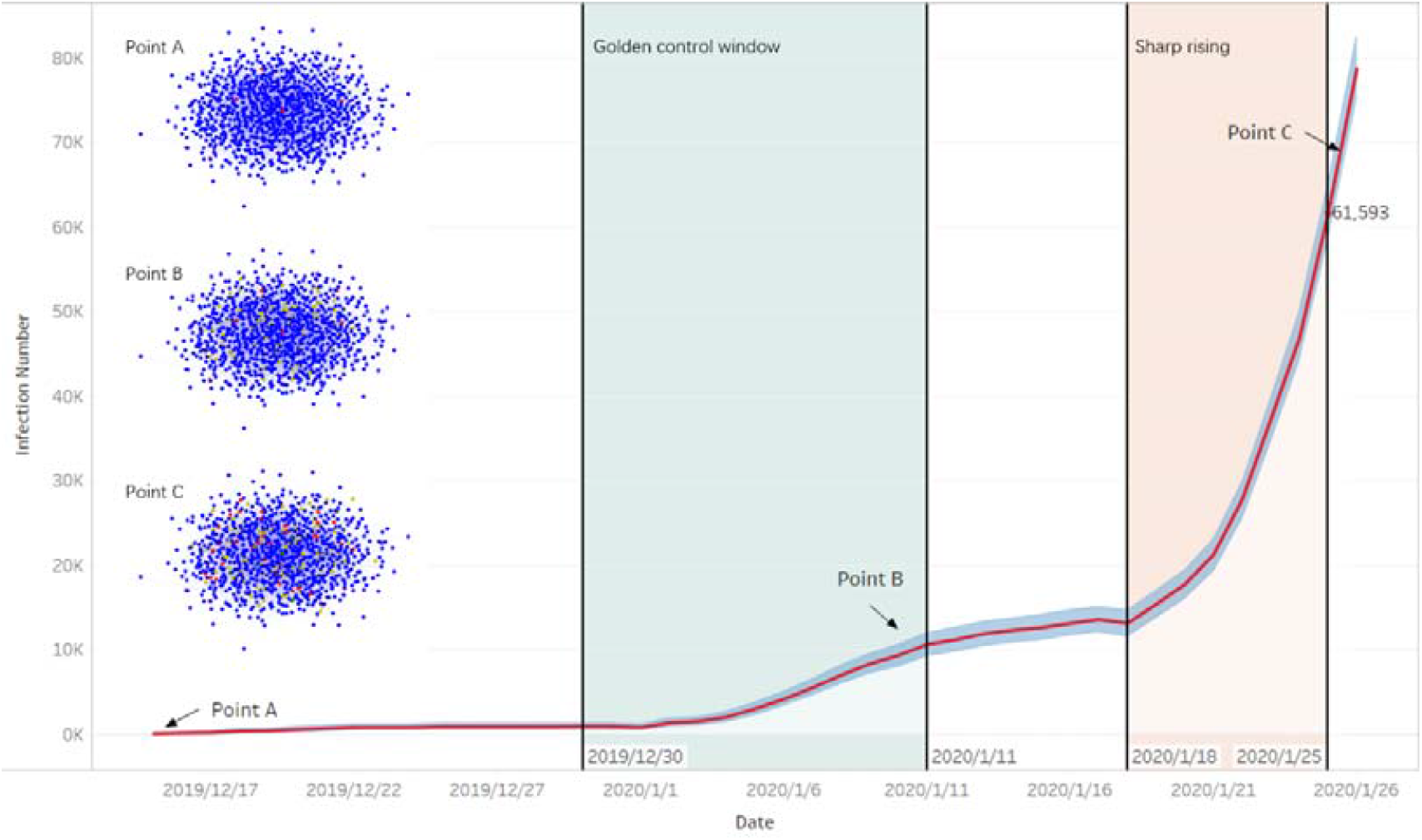
The estimation of infection scale and red line indicate the infection number, while blue band is 95% CI. And the network topology graphs on left (blue: susceptible node, yellow: infected node, red: pathogen node, green: removal node) are simulation of epidemic network development of Wuhan which demonstrate situations of mid-December (point A), January 10(point B) and from 25nd (point C) in Wuhan respectively. It’s noticed that some removal nodes (rehabilitation and death) have occurred in point C.

#### 2. Phrase A and B in Wuhan

During the 1^st^ week after the closure of the city in Wuhan, relative shortage of medical resources and crowd shopping for supplies may induce gathering and cross-infection. Therefore, the contact rate in Phrase A could be considered to be adjusted to 1/15 of the baseline, while the contagious probability is adjusted to 0.5 times the baseline. However, these situations would be relieved after the comprehensive aid and response, so the contact rate between people might drop to 1/30 of the baseline in phrase B, even lower. Nevertheless, under the strict control of the confirmed cases and suspected cases, the transmission efficiency could be going down in further. We assumed 70% confirmed cases and suspected cases were quarantined, and probability of infection also reduced to 25% of baseline by the personal protection. As a result, by the model’s calculation, there would be about 3,375 new infections during phrase A and phrase B in Wuhan. The 95% CI is 2,611.31-4,138.69.

From the estimations in different time periods showed above, the total infection scale in Wuhan is 64971, 95%CI 60,955.33-68,986.67.

### Scale estimation in Guangzhou

It’s known that most of the confirmed cases in Non-Wuhan area had exposure history of Wuhan city, and new year migration had accelerated the spreading. We combine the migration data with the model and use Guangzhou city (provincial capital of Guangdong) as an example to: 1. Evaluate the estimation effect of combine model for Non-Wuhan area; 2. Make a cross validation for Wuhan estimation model, because Wuhan estimation results are used as baseline for combine model.

According to Baidu migration data (Baidu’s Migration Index) analysis, combined with Wuhan January 10, 2020 - January 24, 2020 migration index (city was blockaded on 23^rd^, but there were still people moving out of Wuhan according to the migration index even on 24^th^), and the report that the number of people moved out before the 24^th^ is about 5 million people, we can estimate the number of people moving out daily as Table 2. From January 10, 2020 to January 24, 2020, 24,893 people imported to Guangzhou from Wuhan. Based on the infection rate mentioned above, the estimated number of infections is approximately 170, and 95% CI is 161.27-179.26.

**Table 2.** Estimation of population migration from Wuhan to Guangzhou base on the total migration population from Wuhan between Jan 10 to Jan 24.

|  |  |  |  |  |  |
| --- | --- | --- | --- | --- | --- |
| Date | Emigration<br>Index | Emigration<br>estimation<br>(thousand) | Porportion of<br>Guangzou | Population<br>migration<br>(Wuhan to<br>Guangzhou)<sup>a</sup> | Free<br>movement day<br>for migrant<br>population<sup>b</sup> |
| 2020/1/10 | 6.62 | 298.171 | 0.85% | 2534 | 14 |
| 2020/1/11 | 7.56 | 340.509 | 0.62% | 2111 | 13 |
| 2020/1/12 | 6.22 | 280.154 | 0.61% | 1709 | 12 |
| 2020/1/13 | 5.76 | 259.436 | 0.69% | 1790 | 11 |
| 2020/1/14 | 5.46 | 245.923 | 0.62% | 1525 | 10 |
| 2020/1/15 | 5.91 | 266.192 | 0.53% | 1411 | 9 |
| 2020/1/16 | 6 | 270.245 | 0.53% | 1432 | 8 |
| 2020/1/17 | 6.44 | 290.064 | 0.46% | 1334 | 7 |
| 2020/1/18 | 7.71 | 347.266 | 0.39% | 1354 | 6 |
| 2020/1/19 | 7.41 | 333.753 | 0.44% | 1469 | 5 |
| 2020/1/20 | 8.31 | 374.29 | 0.40% | 1497 | 4 |
| 2020/1/21 | 10.74 | 483.74 | 0.41% | 1983 | 3 |
| 2020/1/22 | 11.84 | 533.285 | 0.37% | 1973 | 2 |
| 2020/1/23 | 11.14 | 501.756 | 0.36% | 1806 | 1 |
| 2020/1/24 | 3.89 | 175.209 | 0.55% | 964 | 0 |
| Average free movement day for migrant population from Wuhan to Guangzhou |  |  |  |  | 7.43* |
| *: if k indicates date sequences, $\frac{\sum_{k=1}^{14} a_k \times b_k}{\sum_{k=1}^{14} a_k}$ | | | | | |

Also, according to Baidu’s migration index, Guangzhou’s exported population was about 11.26 million from Jan 10. Guangzhou’s population is 18 million with 9 million resident population (Guangzhou Statistic Bureau) and 9 million migrant population (Xiao et al.,2018). Therefore, the population base of Guangzhou after January 24 was about 7 million.

According to the timeline, Guangzhou launched level 1 public health response on Jan 23, entered a tight control period and used various protective measures to block transmission. We assume that the contact rate is about 1/10 of the usual (baseline). After Guangzhou started a level 1 public health response, people from Wuhan are checked and asked for self-isolation. In other words, the maximum free movement time of the imported infected cases is 14 days, the minimum is 0 day. By accumulating the free movement time, we could come to the result that the average time window of the activity of the imported infected people is less than 8 days (Table 2). With the assumption above, the total number of infections in Guangzhou after Jan 24 was 315, and 95% CI was 109.20-520.79. Because of the tight control and strict self-quarantine, 315 could be the total infection number.

## Discussion

In this paper, the scale of 2019-nCov infection is simulated and estimated according to the time line of the epidemic at different times based on the methodology of infection dynamics of population topology network. We use historical data and SEIR model for important parameters assumption and these parameters are fitted in simulated infection network. Furthermore, migration data are also used to estimate the area outside Wuhan (Guangzhou is selected here) which could also be regarded as cross-validation materials. The model reproduces a relative comprehensive progression of the infection development in Wuhan, which shows the “golden control window” and a sharp rising of infection scale with time points. And the model also performs well on scale estimation especially in Guangzhou: we estimate the imported cases from Wuhan of Guangzhou is 170 (95%CI: 161.27-179.26) while the number from authority is 168, and we estimated the total infection scale of Guangzhou on Jan 24 is 315 (95%CI: 109.20-520.79) and 339 cases were confirmed in Guangzhou on Feb 18 according to the report of Health Commission of Guangzhou. And it should be noted that the simulation of Guangzhou ia based on the infection scale estimation and the migration data of Wuhan.

Our study shows that dynamic network model and dynamic parameter for different time periods are important to modelling, and challenge is also obvious for parameters’ deduction. Further work should be done for adding more area data for validation especially in the territories outside the mainland of China.

## Data Availability

All data were from the public.

